# Portal Vein Diameter on Routine Clinical CT: Establishing Normals and Disease Associations

**DOI:** 10.64898/2026.02.11.26346009

**Authors:** Katherine Hartmann, Cameron Beeche, Renae Judy, Daniel M. DePietro, Walter R. Witschey, Jeffrey Duda, James Gee, Terence Gade, Penn Medicine Biobank, Michael G. Levin, Scott M. Damrauer

## Abstract

**Purpose:** Portal hypertension, a major complication of chronic liver disease, leads to significant morbidity and mortality. While portal vein diameter measured on imaging has long been proposed as a non-invasive marker of portal hypertension, normative CT-based reference values and population-level associations remain incompletely characterized. Here, we aim to define contemporary reference values for portal vein diameter on clinically obtained CT and evaluate its associations with demographic, clinical, and imaging factors, as well as its diagnostic performance for portal hypertension.

**Methods:** We conducted a retrospective analysis of 20,225 clinically obtained CT scans at a single academic medical center. The main portal vein was automatically segmented using Total Segmentator, and maximum diameter extracted using the Vascular Modeling Toolkit. Associations with demographic and imaging factors were evaluated using linear mixed-effects models; prevalent liver disease and portal hypertension using logistic regression; risk of incident ascites and esophageal varices among participants with liver disease using Cox regression; and invasive hepatic venous pressures using correlation analysis and linear regression.

**Results:** The mean portal vein diameter was 12.4 mm (95% CI, 12.37–12.45). Larger diameter was independently associated with male sex (+1.4 mm), higher BMI (+0.11 mm/kg/m^2^), greater height (+0.04 mm/cm), and older age (+0.05 mm/10 years) (all p *<*0.001), and was substantially larger on contrast-enhanced abdomen/pelvis CT (+2.4 mm, p *<*0.001). Each 1-mm increase in portal vein diameter was associated with higher odds of prevalent liver disease (OR 1.06; 95% CI, 1.04–1.08) and portal hypertension (OR 1.18; 95% CI, 1.12–1.28). Among individuals with liver disease, greater diameter predicted higher risk of incident esophageal varices (baseline diameter HR 1.50; 95% CI, 1.14–2.08) and ascites (HR per mm increase in diameter 1.06; 95% CI, 1.003–1.12). However, portal vein diameter demonstrated weak to no association with invasively measured hepatic venous pressures.

**Conclusion:** In this large, EHR-linked imaging cohort, the mean portal vein diameter on CT was 12.4 mm and varied with demographic and imaging factors. Larger diameter was associated with liver disease, portal hypertension, and subsequent development of varices and ascites, supporting use of portal vein diameter as a pragmatic screening or enrichment tool within multimodal clinical frameworks.

**Key Results:**

1. Mean portal vein diameter on routine clinical CT was 12.4 mm (95% CI, 12.37–12.45) and varied with sex, height, BMI, exam type, contrast use, and clinical setting.
2. Each 1-mm increase in portal vein diameter was associated with higher odds of prevalent liver disease (OR 1.06) and portal hypertension (OR 1.18).
3. Among individuals with liver disease, larger portal vein diameter predicted higher risk of incident esophageal varices and ascites, independent of demographic and imaging factors.

## 1 Introduction

The main portal vein dilates and increases in diameter in response to portal hypertension, which is a major consequence of chronic liver disease. Portal hypertension can lead to serious complications, including refractory ascites and development of portosystemic collateral vessels. Among these, esophageal varices are of particular concern, as rupture may result in catastrophic gastrointestinal bleeding and significant mortality [1]. Early identification of portal hypertension is therefore critical for risk stratification, surveillance, and timely intervention.

The current reference standard for assessing portal pressures is invasive hepatic venous pressure gradient measurement. However, there is interest in identifying reliable, non-invasive markers that could signal early portal hypertension to better guide further diagnostic testing and disease management [2, 3]. Cross-sectional imaging with computed tomography (CT) is routinely obtained for a wide variety of clinical indications, and portal vein diameter, readily measurable on these studies, has long been proposed as a candidate marker of portal hypertension [2, 4–7].

Much of the foundational work defining a “normal” portal vein diameter has been based on ultrasound, with 13 mm frequently cited as the upper limit of normal [1, 2, 8, 9]. However, sonographic measurements are highly operator dependent and influenced by patient positioning and acoustic window limitations. More recent work using CT has suggested that a normal portal vein may be larger, with reported average diameters as high as 15–16 mm [10–12]; yet, CT-based normative values remain incompletely defined.

We evaluated 20,225 CT scans acquired during routine clinical care to establish reference values for portal vein diameter on CT. In addition, we investigated associations between portal vein diameter, imaging acquisition parameters, patient demographics, and clinical variables, including hepatic venous pressures. These data provide an updated framework for interpreting portal vein diameter on CT and assessing its utility as a non-invasive marker of portal hypertension.

## 2 Methods

A retrospective cohort analysis was undertaken to evaluate the maximum portal vein diameter measured on CT scans acquired for routine medical care. This study was approved by the University of Pennsylvania Institutional Review Board (Protocol # 858471).

### Data

#### Main Portal Vein Diameter Measurements

The portal vein was segmented from clinically obtained CT scans from the Penn Medicine Biobank using *Total Segmentator*, a U-Net-based semantic segmentation architecture [13], which automatically segments the portal-splenic vein. We considered only CT scans of the chest with and without contrast and abdomen/pelvis with and without contrast corresponding to CPT codes 71260, 71250, 74177, and 74176. Erosion and dilation morphological operations were used to remove erroneous gaps in the segmentation map. The largest continual region was extracted and a three-dimensional mesh representation of portal-splenic vein was computed using the marching cubes algorithm, followed by Laplacian smoothing. The centerline of the three-dimensional mesh was extracted using VMTK3 from the *Vascular Modeling Toolkit* [14, 15]. The diameter was calculated as the average distance from the centerline to the nearest five points on the three-dimensional mesh representation, multiplied by two. The diameter of the portal vein was considered to be the maximum of these values.

A subset of 100 images were reviewed by a radiologist and the maximum portal vein diameter was assessed manually by taking the maximum portal vein diameter in the axial plane.

#### Splenic Volume Measurements

The spleen was segmented using Total Segmentator, as above. *pyradiomics*, an imaging library implemented in Python, was used to generate a triangular surface mesh and calculate the enclosed 3D volume of that mesh (MeshVolume). The maximum volume across different series in the imaging study was taken as the splenic volume.

#### Clinical Covariates

Age, sex, race, height, and BMI were considered to be potentially relevant clinical co-variates. Age and sex are recorded in the imaging metadata and were obtained using Montage (Nuance Powerscribe, Burlington, MA). Race, Height and BMI were derived from the electronic health record and curated as part of the Penn Medicine Biobank (v3 release) [16]. Race categories with fewer than 10 participants (American Indian or Alaska Native; Native Hawaiian or Other Pacific Islander) were excluded from regression analyses due to insufficient sample size for stable effect estimation; these categories are reported in descriptive summaries only. Height and BMI measurements closest to the date of the CT scan were selected.

#### Hepatic Venous Pressures

Montage was used to gather reports for those individuals in the cohort who had interventional radiology procedures that included hepatic venous pressures. Hepatic wedge pressures and corrected hepatic venous pressure gradients were extracted from radiology reports using text parsing with manual review of 30-50 entries that failed rules based parsing.

#### Portal Hypertension and Liver Disease

Portal hypertension and liver disease were defined using phecodes derived from the electronic health record and curated as part of the Penn Medicine Biobank [17, 18]. Portal hypertension was defined by phecode 571.81. Liver disease was defined using phecodes 155 (cancer of liver and intrahepatic bile ducts), 198.4 (secondary malignancy of liver), 317.11 (alcoholic liver damage), 571 (cirrhosis), 573 (other diseases of liver), and 750.22 (congenital anomaly of gallbladder, bile ducts, liver, pancreas). As falsification endpoints, we also gathered phecodes representing osteoarthritis (740) and lipoma (214.1). We considered phecodes that were present prior to and after the date of the CT scan, corresponding to prevalent and incident disease.

#### Scan Parameters

Relevant scan parameters including presence or absence of intravenous contrast, exam type (e.g. CT chest, CT abdomen/pelvis), and exam site (emergency, inpatient, outpatient) were obtained using Montage.

### Statistical Analysis

A randomly selected subset of 100 CT examinations was independently reviewed by a board-eligible radiologist, and portal vein diameter was manually measured using standard clinical tools. Agreement between automated and manual measurements was assessed using Bland–Altman analysis, the intraclass correlation coefficient (two-way, absolute agreement, single measures), and paired equivalence testing using the two one-sided tests (TOST) procedure with equivalence bounds set at ±12% of the mean manual measurement. This margin was selected to reflect the magnitude of variability reported in prior studies of portal vein diameter measurement on CT [10, 11].

The relationship between portal vein diameter and spleen volume was assessed using Pearson correlation analyses restricted to the first exam from each individual. Spleen volume was modeled as a continuous outcome variable in linear mixed-effects models, including all exams for individuals and incorporating a random intercept to account for participants with multiple examinations.

Portal vein diameter was summarized using mean and standard deviation as well as the 95% confidence interval. Univariable and multivariable linear mixed-effects models were used to evaluate associations between diameter and demographic as well as scan-related variables, again incorporating a random intercept to account for participants with multiple examinations. Differences in mean portal vein diameter across exam types were assessed using pairwise t-tests for the first exam from each individual.

To evaluate whether portal vein diameter associated with clinical outcomes, prevalent liver disease and prevalent portal hypertension (defined prior to the index CT examination) were modeled separately as outcomes with portal vein diameter as the primary explanatory variable. Given that disease status was largely constant within individuals and most participants contributed only one exam, patient-level random-intercept models were unstable. Accordingly, we used logistic regression with cluster-robust standard errors at the participant level, which accounts for within-participant correlation arising from repeated examinations [19, 20].

We next evaluated whether portal vein diameter among patients with prevalent liver disease including cirrhosis was informative of subsequent clinical outcomes, specifically incident esophageal varices and ascites. Time-to-event analyses were performed using Cox proportional hazards regression. Time zero (baseline) was defined as the first CT examination available after a previously established diagnosis of liver disease and prior to any documented diagnosis of ascites or esophageal varices. Participants were followed from baseline until development of the outcome of interest, death or end of follow-up, whichever occurred first. Follow-up was administratively censored on September 23, 2024, corresponding to the last date of available phenotype data. Portal vein diameter was modeled as a continuous predictor with adjustment for demographic and clinical covariates. Hazard ratios (HRs) with 95% confidence intervals were derived from exponentiated model coefficients. The proportional hazards assumption was evaluated using Schoenfeld residuals. For models in which non-proportionality was detected in categorical covariates, we fit stratified Cox models. For models in which non-proportionality was detected in numeric covariates or portal vein diameter, we specified a log(time)-by-variable interaction term tt(Variable) = Variable*×*log(time) to allow the hazard ratio associated with the given variable to vary over follow-up. Time to event curves were generated by dichotomizing portal vein diameter using a prespecified threshold of 13 mm (*>*13 mm vs *≤* 13 mm). Cumulative incidence was estimated using the Kaplan–Meier method and stratified by portal vein diameter category. Event curves were plotted as cumulative incidence functions, with between-group differences assessed using the log-rank test. Follow-up was truncated at 5 years to focus on clinically relevant risk intervals. To assess residual confounding and association specificity, we examined prespecified falsification endpoints, osteoarthritis and lipoma, for which no biologically plausible relationship with portal vein diameter was expected.

To evaluate the relationship between portal vein diameter and the gold standard for portal hypertension assessment, invasive hepatic venous pressure measurements, we analyzed participants who underwent hepatic venous pressure gradient evaluation. For correlation analyses, Pearson correlation coefficients were calculated to assess the linear association between portal vein diameter and both wedged hepatic pressure and hepatic venous pressure gradient, using the hepatic venous measurement closest in time to the CT examination for each participant. For regression analyses, all available paired portal vein and hepatic venous pressure measurements were included. Hepatic venous pressures were modeled as continuous outcome variables with portal vein diameter as the primary explanatory variable and additional demographic and clinical covariates using general linear models with cluster-robust standard errors to account for repeated observations.

## 3 Results

### 3.1 Measurement of Portal Vein Diameter

In 20,225 clinically obtained CT scans representing 15,365 participants, see Table 1, the mean portal vein diameter was 12.4 mm (95% confidence interval 12.38-12.46 mm), see Figure 1. Portal vein diameter was determined using an automated pipeline built from *Total Segmentator*, U-Net-based semantic segmentation architecture [13], which automatically segments the portal-splenic vein and the *Vascular Modeling Toolkit* [15]. In a subset of 100 CT scans that were reviewed manually by a radiologist, the intraclass correlation coefficient between automated and manual measurements was 0.44, (95% confidence interval 0.26-0.58), see Supplementary Figure S1. While the ICC indicated modest to moderate agreement, paired equivalence testing use two onesided tests (TOST) procedure demonstrated that automated and manual portal vein measurements were statistically equivalent within prespecified *±* 12% bounds (mean difference 0.85 mm; overall TOST p-value = 0.008).

**Fig. 1.**
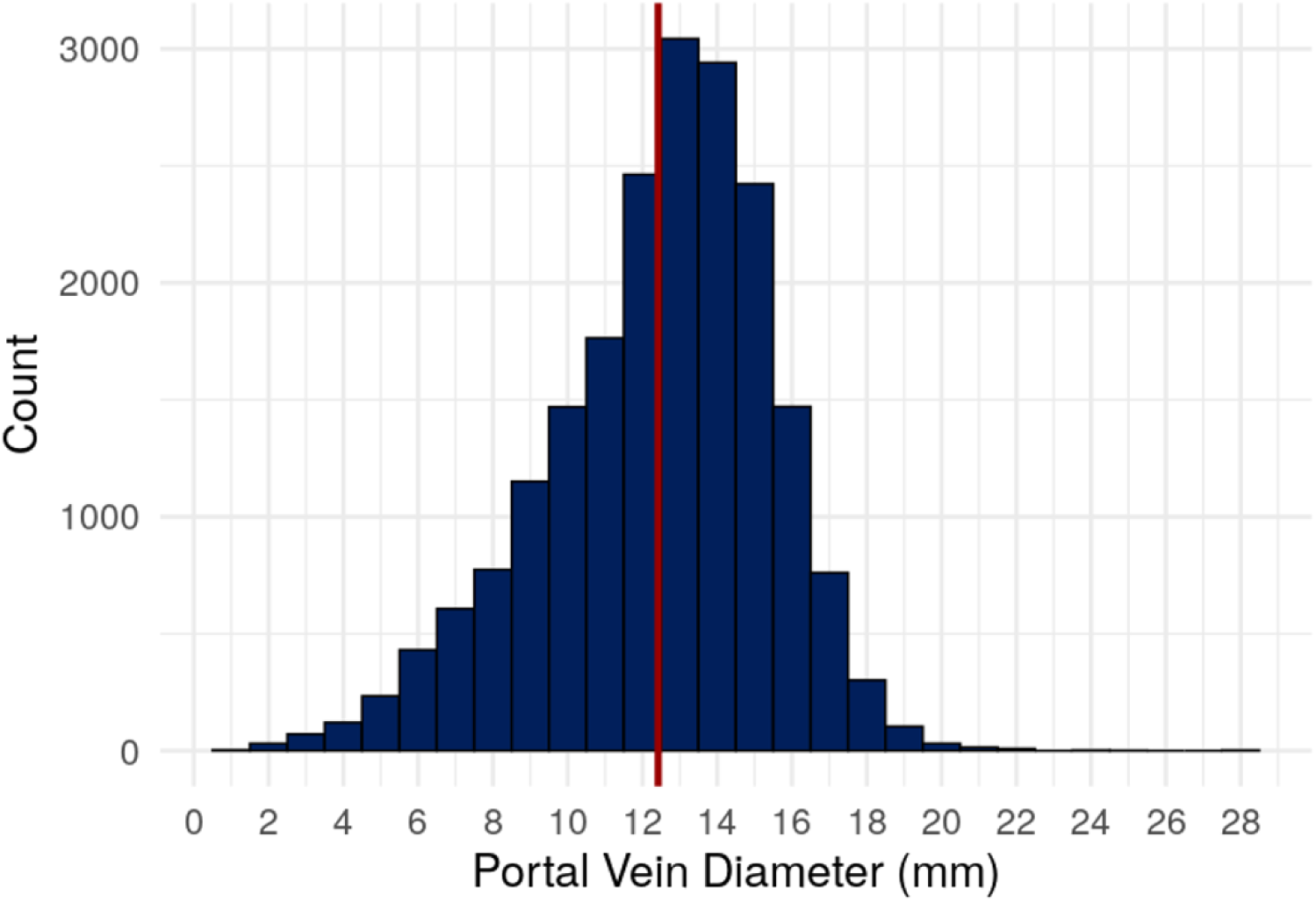
Distribution of maximum portal vein diameter in 20,225 CT scans obtained as part of routine clinical practice.

**Table 1.** Baseline characteristics of the study cohort. summarized at the participant level. For each individual, values reflect person-level summaries from all available CT examinations: sex and race were taken as first recorded; age, height, BMI, and portal vein diameter represent the mean across exams. Liver disease, portal hypertension, osteoarthritis, and lipoma represent prevalent disease, specifically the presence of any diagnosis identified in the electronic health record prior to the date of the CT scan.

| Characteristic | N = 15,365<sup>†</sup> |
| --- | --- |
| Sex |  |
| Female | 7,665 (50%) |
| Male | 7,700 (50%) |
| Race |  |
| American Indian or Alaska Native | <10 (<0.1%) |
| Asian | 235 (1.9%) |
| Black or African American | 3,223 (26%) |
| Native Hawaiian or Other Pacific Islander | <10 (<0.1%) |
| White | 8,389 (68%) |
| More than One Race | 101 (0.8%) |
| Unknown or Not Reported | 419 (3.4%) |
| Age (years) | 60 (15) |
| Height (cm) | 170 (10) |
| BMI (kg/m<sup>2</sup>) | 28 (7) |
| Number of Exams | 1 (1, 14) |
| Liver Disease | 3,958 (26%) |
| Portal Hypertension | 370 (2.4%) |
| Falsification Endpoint: Osteoarthritis | 4,222 (27%) |
| Falsification Endpoint: Lipoma | 220 (1.4%) |

Continuous values are reported as mean (standard deviation) except for *Number of Exams*, which is reported as median (minimum, maximum); categorical values are reported as n (%).

Because splenomegaly is a recognized downstream manifestation of portal hypertension and can be quantified concurrently on the same CT examination as portal vein diameter, we asked whether portal vein diameter was associated with spleen volume. Portal vein diameter showed a positive association with spleen volume (*R*^2^ 0.08; p-value *<*2.2e-16). In univariable linear regression, each 1-mm increase in portal vein diameter corresponded to a 15.3 mL greater spleen volume (95% confidence interval 14.3-16.3 mL; p-value *<*2.2e-16). This association persisted with only modest attenuation after adjustment for demographic (age, BMI, height, sex, race) and scan (site, exam) characteristics (*β* = 10 mL per mm; 95% confidence interval 8.7-11.2 mL; p-value *<*2.2e-16), indicating that larger portal vein diameter is strongly associated with greater spleen volume and suggesting the automated measure of portal vein diameter captures underlying physiology.

### 3.2 Determinants of Portal Vein Diameter

To characterize associations between demographic and scan characteristics with portal vein diameter, we performed regression analysis. Univariable models demonstrated associations with demographic, anthropometric and scan characteristics, see Supplementary Table S1. These associations remained robust in multivariable models, with portal vein diameter independently associated with age, BMI, height, sex, race, site, and exam when additionally controlling for scan timing, see Figure 3 and Supplementary Table S2. For every 10 year increase in age, portal vein diameter increased by 0.06 mm (95% confidence interval 0.025-0.087; p-value 0.0003). For every 1 cm increase in height, portal vein diameter increased by 0.04 mm (95% confidence interval 0.031-0.043 mm; p-value *<*2.2e-16). For every 1kg/m^2^ increase in BMI, maximum portal vein diameter increased by 0.11 mm (95% confidence interval 0.104-0.116 mm; p-value *<*2.2e-16). Portal vein diameter measured on average 1.4 mm greater in men than in women (95% confidence interval 1.23-1.48 mm; p-value *<*2.2e-16). Compared to White participants, portal vein diameter measured 1.3 mm less among participants categorized as Black or African American (95% confidence interval -1.43 to -1.23; p-value *<*2e-16), 0.5 mm less in those categorized as More than One Race (95% confidence interval -0.99 to -0.02; p-value 0.04), and 0.4 mm less in those categorized as Unknown or Not Reported race (95% confidence interval -0.64 to -0.14; p-value 0.002).

Compared to an outpatient setting, portal vein diameter measured 0.8 mm less in emergency department exams (95% confidence interval -0.94 to -0.71; p-value *<*2e- 16) and 0.6 mm less in inpatient exams (95% confidence interval -0.69 to -0.46; p-value *<*2e-16). Compared to CT Abd/Pelvis without contrast, portal vein diameter measured 2.4 mm larger on CT Abd/Pelvis with contrast (95% confidence interval 2.25-2.54; p-value *<*2e-16) and 0.3 mm less on CT chest without contrast (95% confidence interval -0.45 to -0.17; p-value 9e-6), Figure 2.These results suggest that both biological factors and technical acquisition features contribute to variation in measured portal vein diameter.

**Fig. 2.**
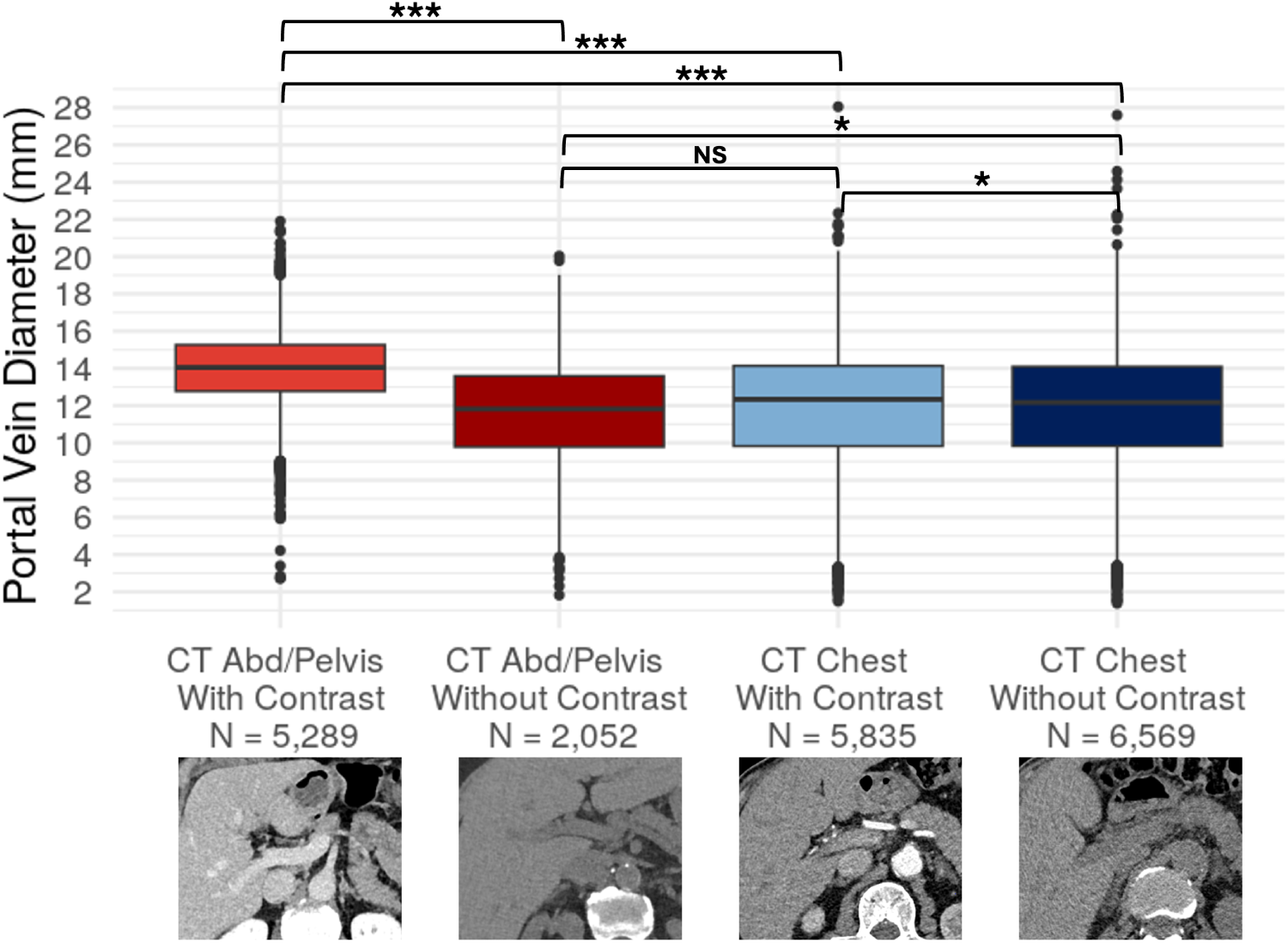
Maximum portal vein diameter by exam type. Boxplot of maximum portal vein diameter (mm) by exam with sample size (N) and representative images from each exam type included below the relevant boxplot. P-values correspond to pairwise t-tests comparing measured portal vein diameters across exam types. NS p-value *>*0.05, * p-value *<*0.0001, *** p-value *<*2.2e-16

**Fig. 3.**
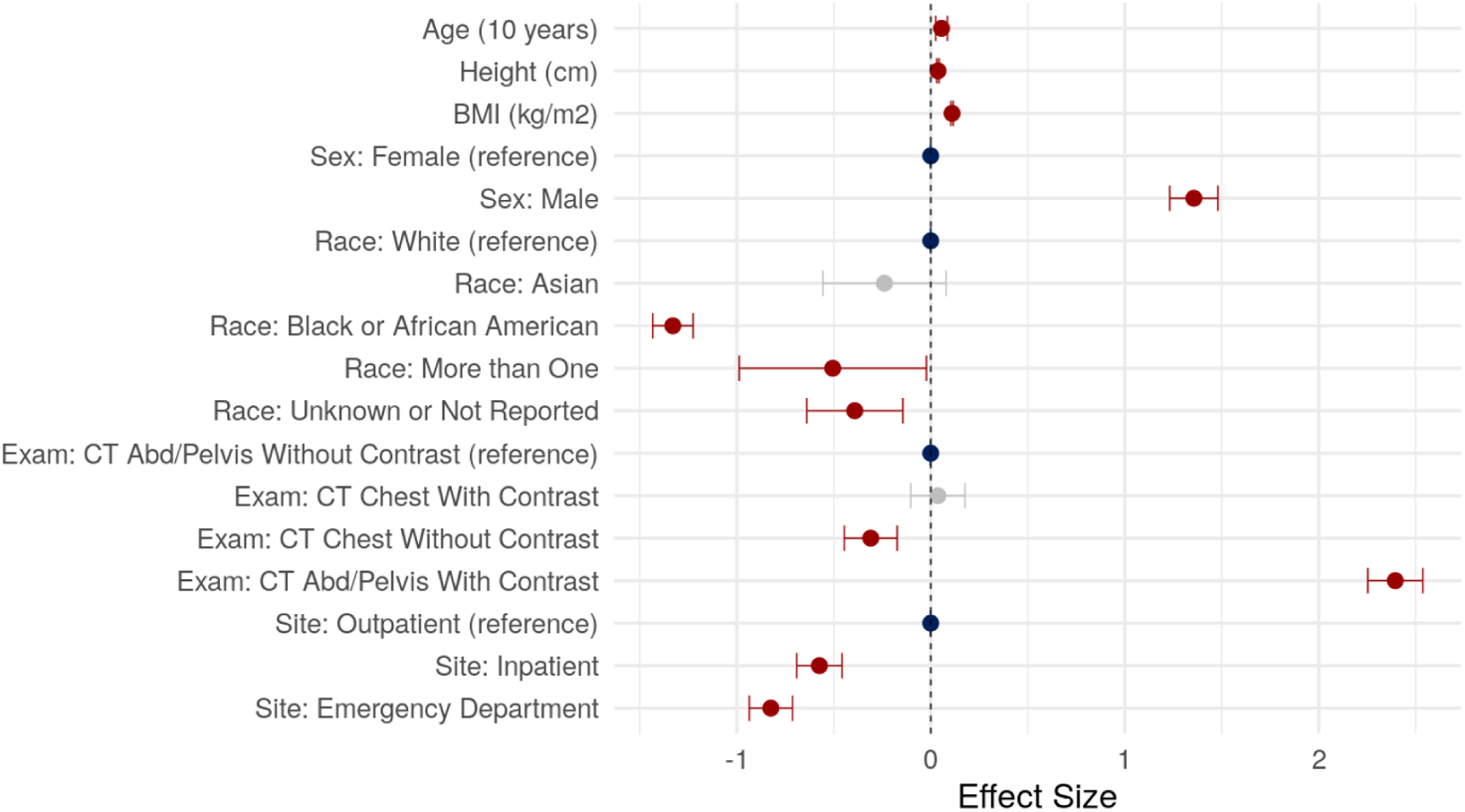
Covariates associated with portal vein diameter in a multivariable mixed-effects model. Adjusted effect sizes for covariates associated with measured portal vein diameter in multivariable linear mixed-effects model. Points represent effect sizes and horizontal lines denote 95% confidence intervals. The vertical dashed line indicates no effect. Variables labeled “reference” and colored in blue represent reference categories. Variables in gray are not statistically significant. All estimates are derived from a fully adjusted multivariable model.

### 3.3 Portal Vein Diameter and Prevalent Liver Disease and Portal Hypertension

Larger portal vein diameter was strongly associated with prevalent liver disease and portal hypertension. These associations were observed in univariable analyses (Table S3) and remained robust in multivariable logistic regression models adjusting for demographic and scan characteristics with participant-clustered robust standard errors. Each 1 mm increase in portal vein diameter was associated with 6% higher odds of prevalent liver disease (OR 1.06; 95% confidence interval 1.04–1.07; p-value 2.5e- 11) and 18% higher odds of prevalent portal hypertension (OR 1.18; 95% confidence interval 1.13–1.22; p-value 6.8e-09), independent of age, height, BMI, sex, race, exam type, and clinical setting. No associations were observed with prespecified falsification endpoints, including osteoarthritis (OR 1.01; 95% confidence interval 0.996–1.02; p- value 0.18) and lipoma (OR 1.035; 95% confidence interval 0.987–1.08; p-value 0.15), supporting specificity for liver-related outcomes, see Figure 4.

**Fig. 4.**
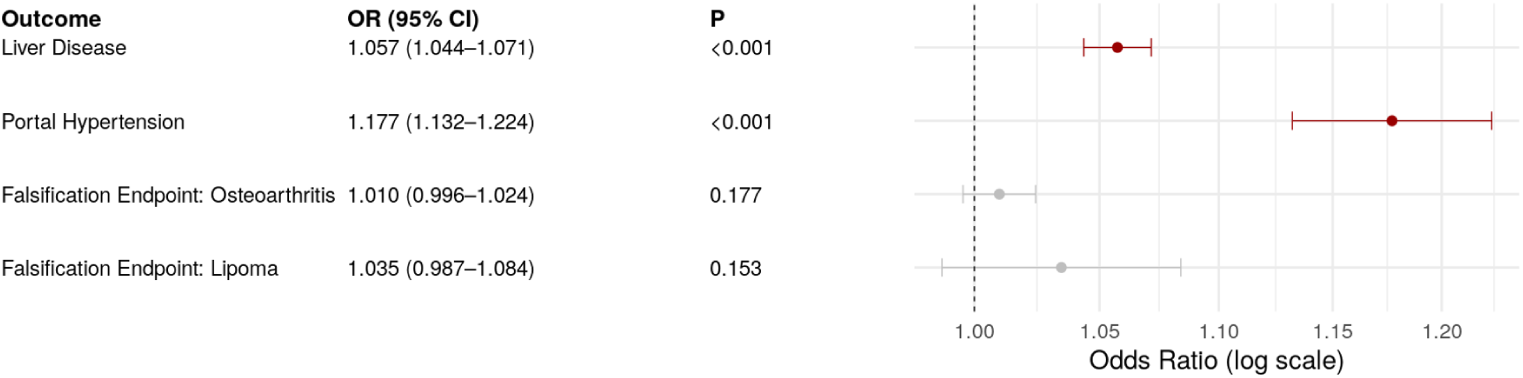
Association of portal vein diameter with prevalent liver disease, portal hypertension, and falsification endpoints. Forest plot showing odds ratios (ORs) per 1 mm increase in portal vein diameter with 95% confidence intervals for associations from multivariable logistic regression with prevalent liver disease, prevalent portal hypertension and falsification endpoints (prevalent osteoarthritis and prevalent lipoma). The vertical dashed line denotes the null value (OR = 1.0).

### 3.4 Portal Vein Diameter and Risk of Incident Esophageal Varices and Ascites

Among participants with prevalent liver disease (Table S4), larger portal vein diameter was associated with higher risk of incident esophageal varices and ascites. This association was observed in univariable analysis (Table S3) and persisted in multivariable models accounting for non-proportional hazards. For esophageal varices, at baseline, each 1 mm increase in portal vein diameter was associated with a 50% higher hazard (HR 1.5; 95% confidence interval 1.14-2.08; p-value 0.005), with attenuation of the association over time. In contrast, for ascites, each 1 mm increase in portal vein diameter was associated with a 6% higher hazard (HR 1.06; 95% confidence interval 1.003–1.12; p-value 0.04), with no evidence of time-varying effects. Kaplan-Meier analysis using a 13-mm threshold demonstrated higher cumulative incidence of varices (5-year cumulative incidence of varices, 4.4% vs 2.7%; log-rank p-value 0.004) and ascites (5-year cumulative incidence of ascites, 8.8% vs 6.9%; log-rank p-value 0.09) among individuals with portal vein diameter *>*13 mm compared with *≤* 13 mm, see Figure 5).

**Fig. 5.**
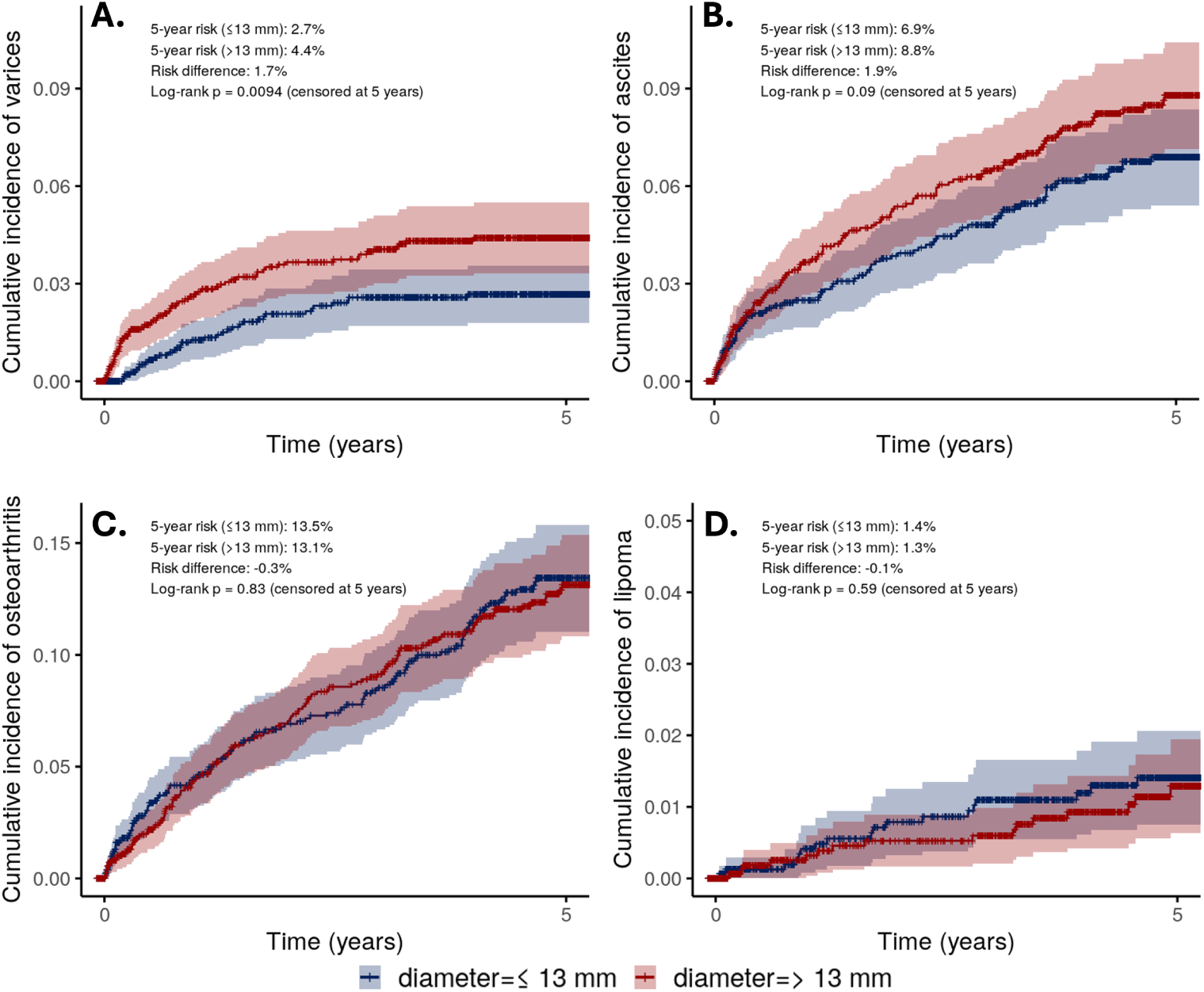
Cumulative incidence of clinical outcomes according to portal vein diameter. Kaplan-Meier curves demonstrate the 5-year cumulative incidence of (A) ascites, (B) esophageal varices, (C) osteoarthritis, and (D) lipoma among participants with prevalent liver disease at baseline, stratified by portal vein diameter *≤* 13 mm (blue) vs *>*13 mm (red) and censored at 5 years. A threshold of 13 mm was used, consistent with prior literature defining portal vein enlargement. Shaded regions represent 95% confidence intervals.

Falsification endpoints were used to assess residual confounding and the specificity of observed associations. Consistent with absence of a biologically plausible relationship, portal vein diameter was not associated with incident osteoarthritis or incident lipoma, with hazard ratios near unity in both univariable (Table S3) and multivariable models (osteoarthritis multivariable adjusted HR 1.01; 95% confidence interval 0.96-1.06; p-value 0.7; lipoma multivariable adjusted HR 1.06; 95% confidence interval 0.94-1.2; p-value 0.3) and Kaplan-Meier analysis without a difference in cumulative incidence among individuals with portal vein diameter *>*13 mm compared with *≤* 13 mm (osteoarthritis 5-year cumulative incidence, 13.5% vs 13.1%; log-rank p-value = 0.83; lipoma 5-year cumulative incidence, 1.4% vs 1.3%; log-rank p-value = 0.59).

### 3.5 Portal Vein Diameter and Invasive Hepatic Venous Pressure Measurements

Among 73 participants who underwent invasive hepatic venous pressure assessment, wedged hepatic pressure and hepatic venous pressure gradients were moderately correlated (r = 0.52; 95% confidence interval, 0.33-0.67; p-value 2e-06), supporting internal physiologic consistency. Measured portal vein diameter was not significantly correlated with either wedged hepatic pressure (r = 0.22; 95% confidence interval, -0.01 to 0.43; p-value 0.07) or hepatic venous pressure gradient (r = 0.2; 95% confidence interval, -0.04 to 0.41; p-value 0.1), see Figure 6. In univariable linear regression, portal vein diameter showed no association with wedged hepatic pressures and was positively associated with hepatic venous pressure gradient, Table S3. However, this was not robust to controlling for demographic and scan related variables including time interval between CT examination and invasive hepatic venous pressure measurement which varied widely (median, 726 days; IQR, 185-1217 days): wedged hepatic pressure *β* = 0.18 mmHg per mm (95% confidence interval -0.22 to 0.58; p-value 0.38) and hepatic venous pressure gradient *β* = 0.06 mmHg per mm (95% confidence interval -0.19 to 0.31; p-value 0.65). Taken together, these results suggest that portal vein diameter does not meaningfully correlate with asynchronously obtained invasive measured portal pressures.

**Fig. 6.**
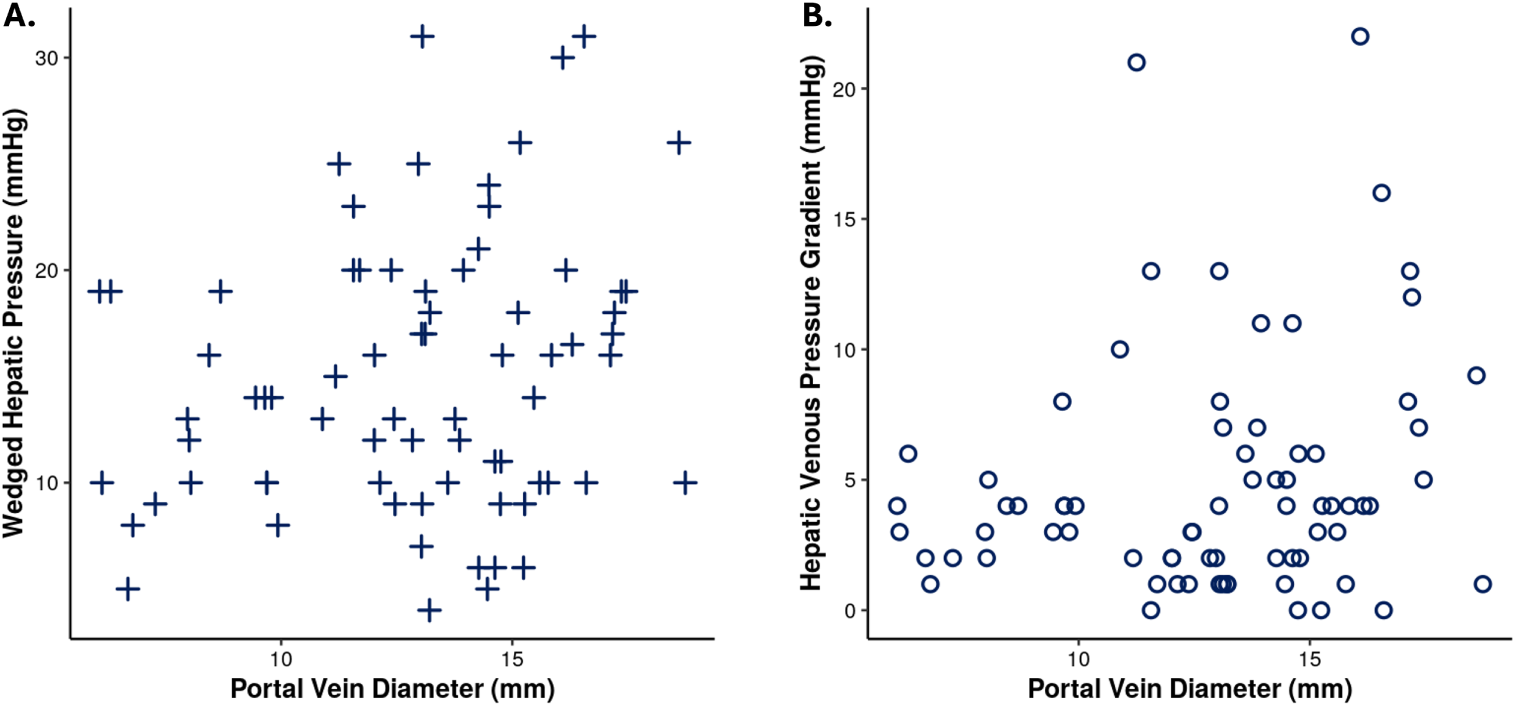
Association between measured portal vein diameter and invasive portal pressure measurements. Scatterplot showing wedged hepatic pressure (A) and hepatic venous pressure gradient (B) as a function of portal vein diameter.

## 4 Discussion

We characterized the distribution of portal vein diameter and assessed its relationships with demographic variables, imaging factors, and liver-related disease phenotypes using a large, real-world imaging cohort. The average maximum portal vein diameter in our population was 12.4 mm (95% confidence interval 12.37-12.45 mm). Portal vein diameter scaled positively with both demographic factors, namely height, BMI, male sex and age, and imaging factors including the type of exam and site of care. Portal vein diameter was associated with prevalent liver disease and portal hypertension and in those with liver disease, incident esophageal varices and ascites. In a limited subsample of individuals with invasive hepatic venous pressure measurements, portal vein diameter did not independently associate with either wedged hepatic pressure or hepatic venous pressure gradient.

In this large, EHR-linked imaging cohort of more than 20,000 clinically acquired CT scans, we show that routinely acquired clinical imaging can be used to empirically determine the distribution of the main portal vein diameter and to characterize its relationship with demographic, imaging and clinical factors. Our estimate of the average maximum portal vein diameter of 12.4 mm falls within previously reported 95% confidence intervals 11.2–19.4 mm albeit slightly lower than prior point estimates of 13.3 mm and 15.5 mm [10, 11]. The 95% confidence interval in our study 12.37–12.45 mm was notably narrower than those previously reported, indicating greater precision attributable to our larger sample size.

Similar to prior studies, we found maximum portal vein diameter was on average larger on CT scans with portal venous contrast [10]. This may reflect blooming artifact from iodinated contrast rather than a true increase in diameter given the relative low volume of fluid given during contrast administration and is an important consideration for pre-procedural and pre-surgical planning prior to portal vein stenting, portal vein reconstructive surgeries, and liver transplantation. Compared to an outpatient setting, portal vein diameter measured 0.8 mm less in emergency department exams and 0.6 mm less in inpatient exams. These differences are more likely explained by variation in patient acuity, volume status and underlying disease burden than by technical artifacts alone.

In those with liver disease, a larger portal vein diameter was associated with subsequent development of clinically relevant portal hypertension outcomes including esophageal varices and ascites. These results suggest that portal vein diameter acquired from routine imaging could function as a pragmatic risk stratification tool, particularly in settings where invasive testing is unavailable, underutilized, or deferred. Whether portal vein diameter can meaningfully complement existing noninvasive assessments of portal hypertension, refine patient selection for surveillance endoscopy, or inform timing of therapeutic intervention remains uncertain. Future studies should evaluate how best to incorporate portal vein diameter into clinical decision frameworks, assess performance alongside established risk markers, and determine whether serial imaging trajectories enhance prognostic precision.

Although portal vein diameter demonstrated consistent and clinically meaningful associations with prevalent liver disease and portal hypertension as well as subsequent development of ascites and esophageal varices, it did not correlate with invasively measured hepatic venous pressures in the subset of individuals having undergone catheter-based assessment. This finding is likely explained by several methodological and biological considerations. First, portal vein diameter, measured on routine clinical CT was obtained at highly variable time points relative to hepatic venous pressures, often separated by years. Given the dynamic nature of portal hypertension and the potential for disease progression, regression, or therapeutic intervention over time, substantial temporal misalignment would be expected to attenuate cross-sectional correlations. Second, the invasive hemodynamic cohort was small, limiting statistical power to detect modest associations and increasing susceptibility to measurement noise. Third, individuals referred for invasive hepatic venous pressure measurement represent a highly selected population with advanced or atypical disease, introducing indication-related selection bias that may further obscure population-level relationships. Even in the absence of concordance with asynchronously obtained invasive pressures, strong and specific associations with clinically relevant outcomes suggest that portal vein diameter captures meaningful portal hypertensive biology.

Our findings should be interpreted in light of several limitations. Disease status was defined using phecodes; although widely used, they may misclassify diagnoses in some cases. Because this was an EHR-based biobank analysis, imaging studies were obtained for clinical rather than population-based indications, introducing the potential for selection and collider bias. We did not observe meaningful associations with falsification endpoints, supporting specificity; however, modest residual confounding cannot be fully excluded in an EHR-based cohort. Finally, the automated measurement approach exhibited only moderate agreement with manual techniques, and despite demonstrated statistical equivalence between automated and manual measurements and correlation with spleen volume suggesting physiologic relevance, thresholds derived from these automated values may not be directly generalizable to routine clinical practice.

Using a large, real-world EHR-linked imaging cohort, we provide a precise, empirically derived estimate of portal vein diameter on routine clinical CT and demonstrate its associations with both demographic and image acquisition features. Despite limited concordance with asynchronously obtained invasive hemodynamic measurements, portal vein diameter was strongly associated with liver disease severity and subsequent development of ascites and esophageal varices, supporting its value as a pragmatic, imaging-derived marker of portal hypertensive biology. These findings highlight the potential of routinely acquired imaging to yield scalable, clinically informative phenotypes while underscoring the need for prospective studies to define how portal vein diameter can be optimally integrated into noninvasive risk stratification and clinical decision-making frameworks.

## Data Availability

Raw data for the analysis dataset are not publicly available to preserve individuals privacy per the Health Insurance Portability and Accountability Act Privacy Rule

## Acknowledgements

We acknowledge the Penn Medicine BioBank (PMBB) for providing data and thank the patient-participants of Penn Medicine who consented to participate in this research program. The PMBB is approved under IRB protocol# 813913 and supported by Perelman School of Medicine at University of Pennsylvania, a gift from the Smilow family, and the National Center for Advancing Translational Sciences of the National Institutes of Health under CTSA award number UL1TR001878.

K.H. was supported by NIH T32 training grant 2-T32-EB-004311-21. WRW was supported by NIH grants R01HL171709-02, R01HL169378-03, P41EB029460-05, R21EB036734-01A1, OT2DOD038048-01.

## 5 Supplement

**Fig. S1.**
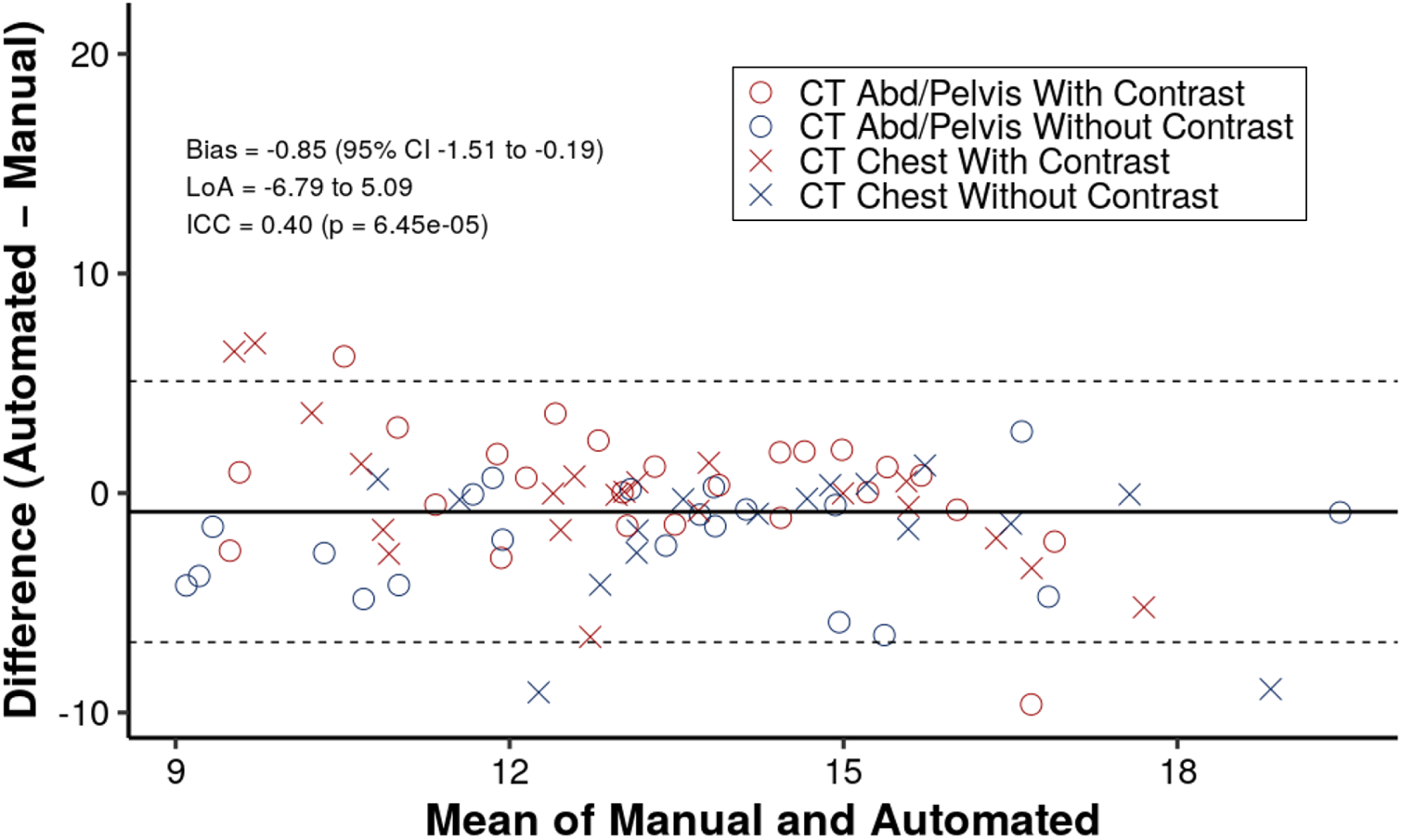
Agreement between automated versus manual measurements. Bland-Altman plot comparing automated and manual portal vein diameter measurements. The x-axis shows the mean of manual and automated measurements, and the y-axis shows their difference (automated - manual). The solid horizontal line indicates the mean bias, and dashed lines indicate the 95% limits of agreement. Points are colored by contrast status (contrast-enhanced vs non-contrast CT) and symbols distinguish exam type (circles = CT Abdomen/Pelvis; crosses = CT Chest). The bias, limits of agreement, and intraclass correlation coefficient (ICC) for absolute agreement is shown.

**Table S1.** Univariable linear mixed-effects models for portal vein diameter.

| Variable | Level | Effect size ($\beta$) | Std. error | p-value |
| --- | --- | --- | --- | --- |
| Age | per 10-years | 0.07 | 0.02 | $2.72 \times 10^{-6}$ |
| Height | per cm | 0.076 | 0.002 | $< 2 \times 10^{-16}$ |
| BMI | per kg/m<sup>2</sup> | 0.069 | 0.004 | $< 2 \times 10^{-16}$ |
| Sex | Female (reference) | — | — | — |
| | Male | 1.737 | 0.045 | $< 2 \times 10^{-16}$ |
| Race | White (reference) | — | — | — |
|  | Asian | -0.711 | 0.192 | 0.00021 |
| | Black or African American | -1.354 | 0.060 | $< 2 \times 10^{-16}$ |
|  | More than One Race | -0.771 | 0.293 | 0.0084 |
|  | Unknown or Not Reported | -0.551 | 0.146 | 0.00017 |
| Exam type | CT Abd/Pelvis Without Contrast (reference) | — | — | — |
| | CT Abd/Pelvis With Contrast | 2.267 | 0.071 | $< 2 \times 10^{-16}$ |
|  | CT Chest With Contrast | 0.132 | 0.070 | 0.06 |
|  | CT Chest Without Contrast | 0.187 | 0.068 | 0.006 |
| Site | Outpatient (reference) | — | — | — |
| | Emergency Department | -0.685 | 0.055 | $< 2 \times 10^{-16}$ |
| | Inpatient | -0.829 | 0.060 | $< 2 \times 10^{-16}$ |
| Year | 2003 (reference) | — | — | — |
|  | 2004 | 2.165 | 3.690 | 0.56 |
|  | 2005 | 2.646 | 3.013 | 0.38 |
|  | 2006 | 0.644 | 2.416 | 0.79 |
|  | 2007 | -0.547 | 2.218 | 0.81 |
|  | 2008 | -0.822 | 2.192 | 0.71 |
|  | 2009 | -0.398 | 2.142 | 0.85 |
|  | 2010 | -0.643 | 2.141 | 0.76 |
|  | 2011 | -0.177 | 2.137 | 0.93 |
|  | 2012 | -0.055 | 2.136 | 0.98 |
|  | 2013 | 0.152 | 2.139 | 0.94 |
|  | 2014 | -0.145 | 2.137 | 0.95 |
|  | 2015 | 0.564 | 2.133 | 0.79 |
|  | 2016 | 0.416 | 2.133 | 0.85 |
|  | 2017 | 0.437 | 2.132 | 0.84 |
|  | 2018 | 0.548 | 2.132 | 0.8 |
|  | 2019 | 1.055 | 2.131 | 0.62 |
|  | 2020 | 1.045 | 2.131 | 0.62 |
|  | 2021 | 1.008 | 2.131 | 0.64 |
| Month | 01 (reference) | — | — | — |
|  | 02 | -0.067 | 0.102 | 0.51 |
|  | 03 | -0.072 | 0.107 | 0.50 |
|  | 04 | -0.207 | 0.103 | 0.05 |
|  | 05 | -0.038 | 0.104 | 0.72 |
|  | 06 | -0.113 | 0.094 | 0.23 |
|  | 07 | -0.099 | 0.096 | 0.3 |
|  | 08 | -0.122 | 0.094 | 0.19 |
|  | 09 | -0.039 | 0.100 | 0.7 |
|  | 10 | -0.054 | 0.090 | 0.55 |
|  | 11 | 0.033 | 0.093 | 0.72 |
|  | 12 | -0.058 | 0.096 | 0.55 |

**Table S2.** Multivariable linear mixed-effects model for portal vein diameter.

| Variable | Level | Effect size ($\beta$) | Std. error | p-value |
| --- | --- | --- | --- | --- |
| Age | per 10-years | 0.056 | 0.016 | 0.00034 |
| Height | per cm | 0.037 | 0.003 | $< 2 \times 10^{-16}$ |
| BMI | per kg/m<sup>2</sup> | 0.110 | 0.003 | $< 2 \times 10^{-16}$ |
| Sex | Female (reference) | — | — | — |
| | Male | 1.356 | 0.063 | $< 2 \times 10^{-16}$ |
| Race | White (reference) | — | — | — |
|  | Asian | -0.239 | 0.162 | 0.14 |
| | Black or African American | -1.329 | 0.053 | $< 2 \times 10^{-16}$ |
|  | More than One Race | -0.505 | 0.246 | 0.04 |
|  | Unknown or Not Reported | -0.392 | 0.126 | 0.002 |
| Site | Outpatient (reference) | — | — | — |
| | Emergency | -0.824 | 0.057 | $< 2 \times 10^{-16}$ |
| | Inpatient | -0.574 | 0.060 | $< 2 \times 10^{-16}$ |
| Exam | CT Abd/Pelvis Without Contrast (reference) | — | — | — |
| | CT Abd/Pelvis With Contrast | 2.395 | 0.072 | $< 2 \times 10^{-16}$ |
|  | CT Chest With Contrast | 0.037 | 0.071 | 0.61 |
| | CT Chest Without Contrast | -0.310 | 0.070 | $8.63 \times 10^{-6}$ |
| Year | 2003 (reference) | — | — | — |
|  | 2004 | 3.293 | 3.085 | 0.29 |
|  | 2005 | 2.263 | 2.519 | 0.37 |
|  | 2006 | 0.079 | 2.107 | 0.97 |
|  | 2007 | -0.145 | 1.873 | 0.94 |
|  | 2008 | -0.724 | 1.854 | 0.70 |
|  | 2009 | -0.120 | 1.798 | 0.95 |
|  | 2010 | -0.140 | 1.795 | 0.94 |
|  | 2011 | 0.307 | 1.788 | 0.86 |
|  | 2012 | 0.238 | 1.787 | 0.89 |
|  | 2013 | 0.672 | 1.789 | 0.71 |
|  | 2014 | 0.441 | 1.787 | 0.81 |
|  | 2015 | 0.599 | 1.784 | 0.74 |
|  | 2016 | 0.689 | 1.783 | 0.70 |
|  | 2017 | 0.797 | 1.783 | 0.65 |
|  | 2018 | 0.877 | 1.783 | 0.62 |
|  | 2019 | 1.333 | 1.782 | 0.45 |
|  | 2020 | 1.391 | 1.782 | 0.43 |
|  | 2021 | 1.320 | 1.782 | 0.46 |
| Month | 01 (reference) | — | — | — |
|  | 02 | -0.006 | 0.097 | 0.95 |
|  | 03 | 0.001 | 0.102 | 0.99 |
|  | 04 | -0.006 | 0.101 | 0.95 |
|  | 05 | 0.076 | 0.100 | 0.45 |
|  | 06 | 0.019 | 0.092 | 0.84 |
|  | 07 | -0.019 | 0.095 | 0.84 |
|  | 08 | 0.013 | 0.092 | 0.89 |
|  | 09 | 0.087 | 0.098 | 0.37 |
|  | 10 | 0.089 | 0.090 | 0.32 |
|  | 11 | 0.061 | 0.093 | 0.51 |
|  | 12 | 0.015 | 0.097 | 0.88 |

**Table S3.**
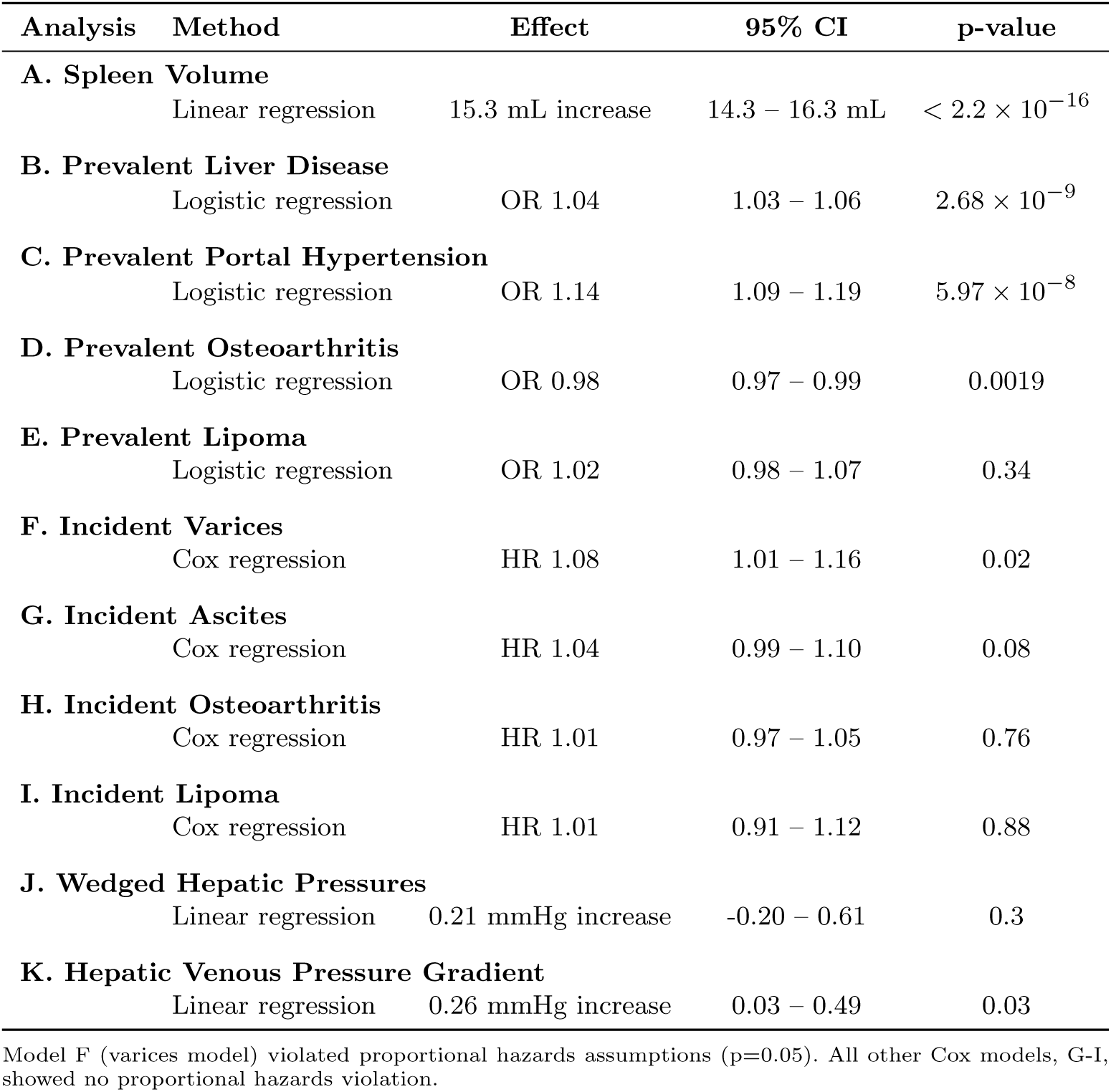
Univariable associations per 1-mm increase in portal vein diameter with various clinical outcomes.

**Table S4.** Baseline Characteristics of Participants with Prevalent Liver Disease. . For each participant, values reflect person-level summaries from all available CT examinations: sex and race were taken as first recorded; age, height, BMI, and portal vein diameter represent the mean across examinations. Ascites, varices, osteoarthritis, and lipoma represent incident disease, specifically the first diagnosis in the electronic health record appearing subsequent to the date of the CT scan.

| Characteristic | N = 3,958<sup>†</sup> |
| --- | --- |
| Sex |  |
| Female | 1,774 (45%) |
| Male | 2,184 (55%) |
| Race |  |
| American Indian or Alaska Native | 1 (<0.1%) |
| Asian | 74 (1.9%) |
| Black or African American | 1,071 (27%) |
| Native Hawaiian or Other Pacific Islander | 2 (<0.1%) |
| White | 2,645 (67%) |
| More than One Race | 27 (0.7%) |
| Unknown or Not Reported | 138 (3.5%) |
| Age (years) | 61 (14) |
| Height (cm) | 170 (10) |
| BMI (kg/m<sup>2</sup>) | 28 (7) |
| Number of Exams | 1 (1, 14) |
| Incident – Ascites | 212 (5.4%) |
| Incident – Varices | 103 (2.6%) |
| Incident – Osteoarthritis | 500 (13%) |
| Incident – Lipoma | 37 (0.9%) |

<sup>†</sup> Continuous values are reported as mean (standard deviation) except for *Number of Exams*, which is reported as median (minimum, maximum); categorical values are reported as n (%).

## Supplemental Note: Penn Medicine Biobank Banner Author List and Contribution Statements

### PMBB Leadership Team

Daniel J. Rader, M.D.; Marylyn D. Ritchie, Ph.D.

**Contribution:** All authors contributed to securing funding, study design, and oversight. All authors reviewed the final version of the manuscript.

### Patient Recruitment and Regulatory Oversight

JoEllen Weaver; Nawar Naseer, Ph.D., M.P.H.; Giorgio Sirugo, M.D., Ph.D.; Afiya Poindexter; Jenna Dever; Aidan Harvey; Sydney Linn; Naman Srivastava

**Contributions:** JW manages participant recruitment and regulatory oversight. NN manages participant engagement, assists with regulatory oversight, and supports researcher access. GS supports researcher access. AP, JD, AH, SL, and NS perform recruitment and enrollment of participants.

### Lab Operations

JoEllen Weaver; Meghan Livingstone; Fred Vadivieso; Stephanie DerOhannessian; Teo Tran; Julia Stephanowski; Salma Santos; Ned Haubein, Ph.D.; Joseph Dunn

**Contribution:** JW, ML, FV, and SD oversee lab operations. ML, FV, AK, SD, TT, JS, and SS perform sample processing. NH and JD manage sample tracking and the laboratory information management system.

### Clinical Informatics

Anurag Verma, Ph.D.; Colleen Morse Kripke, M.S., DPT, MSA; Marjorie Risman, M.S.; Renae Judy, B.S.; Colin Wollack, M.S.

**Contribution:** All authors contributed to the development and validation of clinical phenotypes used to identify study participants and (when applicable) controls.

### Genome Informatics

Anurag Verma, Ph.D.; Shefali S. Verma, Ph.D.; Scott Damrauer, M.D.; Yuki Bradford, M.S.; Scott Dudek, M.S.; Theodore Drivas, M.D., Ph.D.; Zachary Rodriguez, Ph.D.

**Contribution:** AV, SSV, and SD developed the analysis, design, and infrastructure needed for quality control of genotype and exome data. YB performed analyses. TD and AV provided variant and gene annotations and interpreted variant function.

